# Intermediate repeat alleles at polyglutamine repeat loci in the Indian population

**DOI:** 10.1101/2025.11.22.25340801

**Authors:** Meghana Janardhanan, Akshayaa Ganesh, Sowmya Devatha Venkatesh, YM Jeevan, Swarna Buddha Nayok, Pavithra Jayasankar, Vikram Holla, Nitish Kamble, Biju Viswanath, Ravi Yadav, Pramod Pal, Sanjeev Jain, Meera Purushottam

**Affiliations:** Molecular Genetics Laboratory, Department of Psychiatry, National Institute of Mental Health and Neurosciences, Bangalore, India; Molecular Genetics Laboratory, Department of Psychiatry, National Institute of Mental Health and Neurosciences, Bangalore (NIMHANS), India; Department of Neurology, National Institute of Mental Health & Neurosciences (NIMHANS), Hosur Road, Bangalore

**Keywords:** Trinucleotide repeats, repeat instability, Somatic mosaicism, intermediate alleles

## Abstract

An expansion of trinucleotide repeats beyond a threshold, at particular loci, is linked to several diseases. The prevalence of these expansion-linked disorders varies across populations, and may be influenced by the prevalence of sub-threshold intermediate alleles, which are prone to expand. The effect of somatic instability of these repeats, adds further complexity to the role of intermediate repeats. We studied the prevalence of intermediate CAG repeat alleles at *SCA1, SCA2, SCA3* and *HTT* in several hundred individuals. These included patients with clinical suspicion of movement disorder (N∼725), those with severe mental illness (N∼ 420), and healthy individuals from the general population (N∼300). Intermediate alleles were defined as those in the range of 36 -38 CAG repeats for *ATXN1*, 45 -59 CAG repeats for *ATXN3*, 27 - 35 CAG repeats for *HTT*, and 27-32 CAG repeats for *ATXN2*. In those with neuromotor symptoms, we found intermediate alleles in 26 out of 561 (4.6 %) individuals genotyped at *ATXN1*, 10 out of 437 (2.2%) individuals at *ATXN3*, 16 out of 686 (2.3%) individuals at *HTT* and 1 out of 484(0.2%) individuals at *ATXN2*. In individuals without a diagnosis of movement disorder (patients with SMI and healthy controls), intermediate alleles were detected in 19 out of 725 (2.6%) individuals at *ATXN1* and 14 out of 687 (2.0%) individuals at *HTT* loci. No intermediate alleles were detected at the *ATXN2* and *ATXN3* loci. The impact of these intermediate alleles on risk of disease in future generations, and prevalence of disease, needs to be evaluated.

## Introduction

Tandem repeats are common, multi-allelic, highly mutable, and polymorphic sequences in the genome. Expansion of CAG trinucleotide repeats is linked to neurodegenerative syndromes, including Huntington’s disease (HD; Huntingtin *HTT*), Spinocerebellar Ataxia type 1 (SCA1; Ataxin-1, *ATXN1*), SCA2 (*ATXN2*), and SCA3 (*ATXN3*). These disorders are inherited in an autosomal dominant pattern, and often show anticipation which is perhaps driven by instability and expansion of the repeat sequence (1).

Their prevalence varies significantly across populations. HD is more frequent in the European population (4.5-8.9 per 100,000), than in Africa (0.02-2.6 per 100,000) and varies considerably in the South American population (2). This variation in prevalence may be influenced by the average CAG repeat length and frequency of different *HTT* gene haplotypes in the general population (3). According to de Mattei et al., (2023), SCA1 accounts for a significant portion of ataxia syndromes in some parts of Europe; Greece (12%), France (14.1–17.4%), Germany (9%), Finland (4%), the Netherlands (6.2%), and Portugal (2.2%), while in Italy the prevalence varied with geographic origin of probands (4). Elsewhere, SCA1 had a lower relative frequency, being absent or infrequent (0-2%) in United Kingdom, Czech Republic, and Norway, but detected in approximately 5% in Spain. In the case of SCA2, France, Spain and Italy, Eastern European countries, such as Serbia (13%), Poland (11.3%), and Czech Republic (11%), exhibit a relatively high prevalence. SCA3 is the most common spinocerebellar ataxia (SCA) worldwide and is detected in 20-50% of families with SCA. It is the most common SCA in Portugal (58-74%), Brazil (69-92%), China (48-49%), the Netherlands (44%), Germany (42%), and Japan (28-63%) among tested SCA cases, but is quite rare in Italy (1%), and South Africa (4%) (5). In Brazil, its high frequency is particularly notable in regions with strong Portuguese ancestry and remains restricted to a few haplotypes when encountered in other parts of the world (6,7). In India too, SCA1 and SCA2 are more common, while SCA3 is more frequent in areas with Portuguese or European admixture (8,9)

Intermediate alleles (IA) are unstable repeats that usually do not cause disease but may expand, while pathogenic repeat alleles manifest clinically. Since these alleles are inherently unstable, large IAs in the population may act as a reservoir for pathogenic expansions in a subsequent generation, and contribute to the varying prevalence as described earlier. These IAs are observed in a significant proportion of the general population, often at *ATXN1* and *HTT* loci (10). In a systematic literature review, IAs in the *HTT* region ranged from 0.45% - 8.7% in the general population, and 0.05% - 5.1% in individuals with a family history of HD (11). Specific haplogroups (A1 and A2 variants) associated with European HD populations are also detected in those with IAs (12). This suggests that IAs may be more likely to expand upon transmission in particular haplogroups.

IAs might also be linked to the risk of other psychiatric and neurological symptoms. Intermediate alleles at the *HTT* locus were more frequent in those diagnosed with Multi-System Atrophy (8.8%) when compared to controls (3.9%) (13). IAs in *HTT, ATXN1*, and *ATXN2* loci also seem to increase the risk of Alzheimer’s Disease (AD), and Fronto-Temporal Dementia (FTD)(14). Low-penetrance *HTT* repeat expansions were detected in 0.3% of Parkinson’s Disease patients and 0.19% of healthy controls (13). IAs may also have a role in the risk of polymorphic symptoms and severe mental illness (SMI), like schizophrenia and mood disorders (15–17). To examine these issues further, there is a need to carry out a systematic evaluation of IAs in the general population, and in individuals with SMI.

We studied the prevalence of IAs at *ATXN1, ATXN2* and *ATXN3* and *HTT* loci in patients with clinical evidence of movement disorder, and also in patients with severe mental illness, as well as individuals from the general population.

## Methodology

We describe the occurrence of IAs in two sets of samples. The first set included individuals referred to the Genetic Counselling and Testing (GCAT) Clinic, with a suspected diagnosis of movement disorder (ataxia or HD, Group1) (N∼686). The other sets had persons with no symptoms of ataxia or movement disorder, population controls (N∼300, Group 2), and those with a severe mental illness (SMI) (N∼420, Group 3) (either schizophrenia or bipolar disorder, diagnosed according to ICD 10). The size of the CAG repeats at *HTT, ATXN1, ATXN2* and *ATXN3* loci was determined following standard protocols (Venkatesh et al., 2018). The Institutional Ethics Committee approved the study and all patients gave written informed consent for the study. The gnomADv3.1.2 which has STR data for the general population was examined for IAs at the four loci studied, in other world populations of interest (Supplementary table 1a and 1b). (https://gnomad.broadinstitute.org/short-tandem-repeats?dataset=gnomad_r3)

## Results

In group1 we detected 53 chromosomes, with an intermediate repeat size in at least one of the genes that were analyzed. These included IAs in 26/561(4.6%) individuals at *ATXN1*, 1/484(0.2%) individual at *ATXN2*, 10/437(2.3%) individuals at *ATXN3*, and 16/686 (2.3%) individuals at the *HTT* locus (Table 1). In a few instances of those with neuromotor symptoms, (3 cases out of 561, in SCA1; 4 out of 437, in SCA3; 5 out of 686, in HD) an IA was detected along with an expanded allele, at the same locus.

**Table 1:**
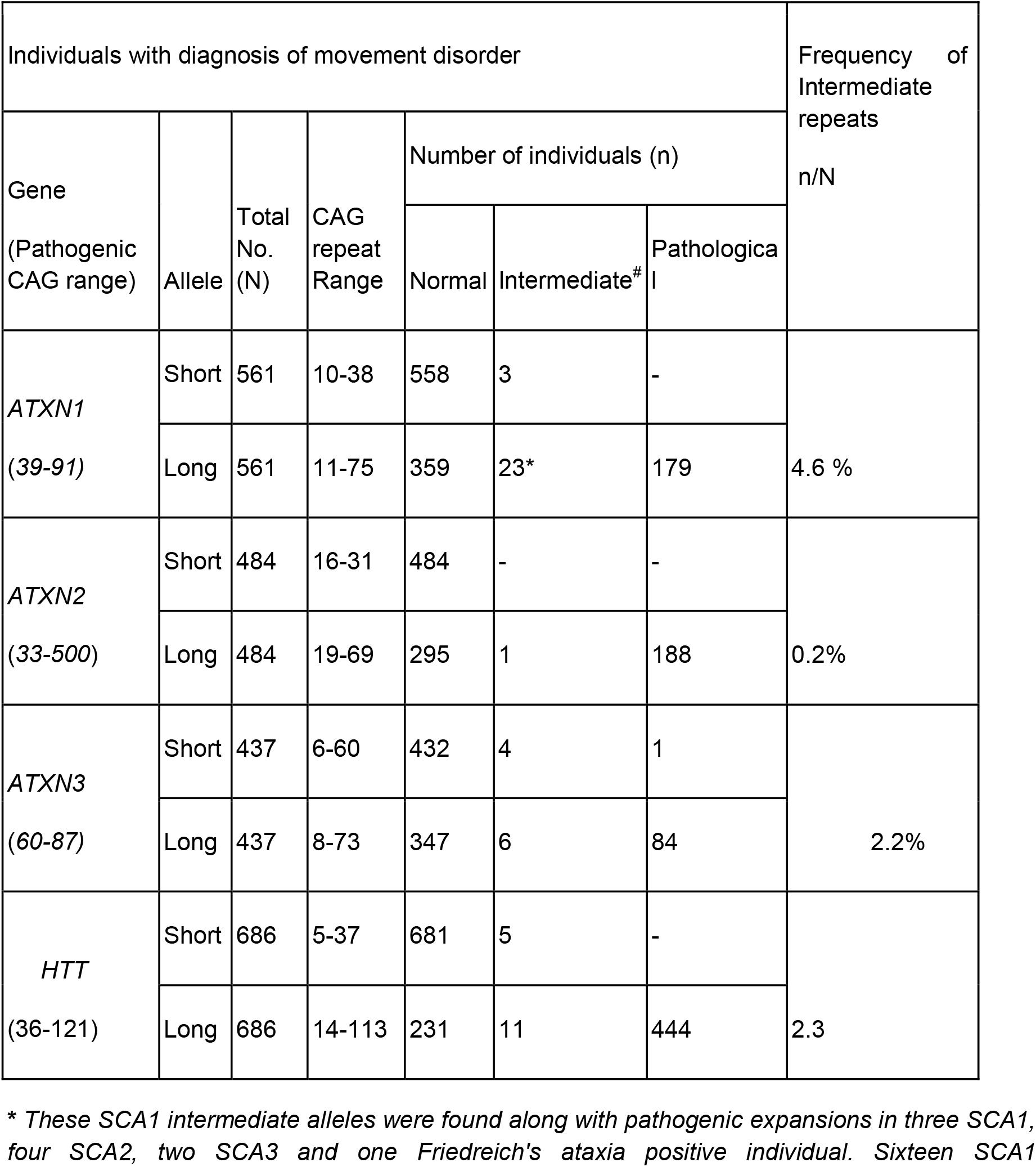
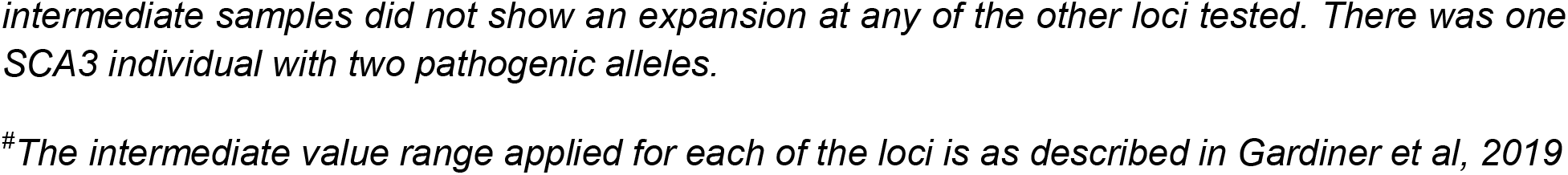
Number of individuals within the normal, intermediate and pathological ranges in individuals with a suspected diagnosis of movement disorder.

| Individuals with diagnosis of movement disorder |  |  |  |  |  |  | Frequency of Intermediate repeats<br>n/N |
| --- | --- | --- | --- | --- | --- | --- | --- |
| Gene<br>(Pathogenic CAG range) | Allele | Total No. (N) | CAG repeat Range | Number of individuals (n) |  |  |  |
|  |  |  |  | Normal | Intermediate<sup>#</sup> | Pathological |  |
| *ATXN1*<br>(39-91) | Short | 561 | 10-38 | 558 | 3 | - | 4.6 % |
|  | Long | 561 | 11-75 | 359 | 23* | 179 |  |
| *ATXN2*<br>(33-500) | Short | 484 | 16-31 | 484 | - | - | 0.2% |
|  | Long | 484 | 19-69 | 295 | 1 | 188 |  |
| *ATXN3*<br>(60-87) | Short | 437 | 6-60 | 432 | 4 | 1 | 2.2% |
|  | Long | 437 | 8-73 | 347 | 6 | 84 |  |
| *HTT*<br>(36-121) | Short | 686 | 5-37 | 681 | 5 | - | 2.3 |
|  | Long | 686 | 14-113 | 231 | 11 | 444 |  |

*\* These SCA1 intermediate alleles were found along with pathogenic expansions in three SCA1, four SCA2, two SCA3 and one Friedreich's ataxia positive individual. Sixteen SCA1*
intermediate samples did not show an expansion at any of the other loci tested. There was one SCA3 individual with two pathogenic alleles.
<sup>#</sup>The intermediate value range applied for each of the loci is as described in Gardiner et al, 2019

In group 2 and 3, IAs were detected in 19/725 (2.6%) individuals (9/308 population controls and 10/417 SMI) at *ATXN1*. Similarly, 14/687(2.0%) individuals (6/302 population controls and 8/377patients with SMI) at *HTT* locus, carried IAs (Table 2). No IAs were observed at the *ATXN2* and *ATXN3* loci.

**Table 2:** Number of individuals within the normal, intermediate and pathological ranges in individuals without a primary diagnosis of ataxia or movement disorder (population controls and individuals with severe mental illness)

| CAG-expansion<br>Disorder<br>Locus | Total<br>Number<br>tested | Number of Individuals |  |  | Prevalence<br>(%) |
| --- | --- | --- | --- | --- | --- |
|  |  | Normal<br>range | Intermediate<br>range | Pathological<br>range |  |
| *ATXN1* | 725 | 706 | 19 | - | 2.6% |
| *ATXN2* | 749 | 749 | - | - | 0 |
| *ATXN3* | 736 | 736 | - | - | 0 |
| *HTT* | 687 | 672 | 14 | 1 | 2.0% |

In summary, IAs were observed in 15/300 controls (4.9%) in group 2, and in group 3, we detected 18 persons with IAs (4.3%). The occurrence of IA did not differ between those with SMI and controls. Moreover, we did not detect any correlation between the age at onset in bipolar disorder (n=187), and CAG upper repeat alleles at any locus.

An examination of population wise STR frequencies in the gnomADv3.1.2 also showed negligible occurrence of IAs at the *ATXN2* and *ATXN3* loci (Supplementary table 1a). IAs are detected in *ATXN1* and *HTT* in about 5% of the global population. At the *ATXN1* locus the IA are seen in less than 1% of those of mid-eastern origin, but in almost 6% of non-Finnish Europeans and 3.5% in the SAS population. At the *HTT* locus too, IAs seem to be undetected in the Amish, but occur in about 5-6% Europeans, and in more than 10% of mid-Eastern samples. (Supplementary table 1b).

## Discussion

A significant proportion of the general population (3-5%) seems to harbor intermediate alleles which seems to be relatively common. Intermediate alleles at the *ATXN1* were also found along with a pathogenic expansion in four SCA2, and two SCA3 positive individuals. Expansions of alleles on both chromosomes have been detected occasionally, in our laboratory in those with ataxia or HD (20) and at other centers (9), and may contribute to more severe disease. Such instances suggest a more generalized defect in DNA replication, leading to slippage of repeat sequences during cell division, perhaps contributing to the occurrence of instability at multiple loci. Previous studies on brain, sperm and blood tissues have also reported somatic instability in both SCA1 and HD, invoking a common mechanism for tissue-specific allele heterogeneity (18). Recent work from our center has also reported regional CAG repeat instability in the *PPP2R2B* locus in samples of postmortem SCA12 brain (19). In a few HD patients and in some siblings and parents of HD patients we found instances of intermediate alleles (20). The presence of IA in DNA from leucocytes could sometimes correspond to pathogenic repeat sizes in other tissues (such as the brain), and thus account for some of the polymorphic phenotypes reported for IAs (17). The *ATXN2* CAG allele in the normal population is almost mono-allelic to a very large extent, suggesting that the repeat length at this locus is quite invariable. At the *ATXN3* locus, the specific haplotype associations suggest founder effects. We did not find IAs in the *ATXN2* and *ATXN3* loci in the non-ataxia population. Associated haplotypes might include sequences that provide structural stability and prevent slippage. Particular DNA sequences might also attract proteins that provide stability. For instance, a unique 17bp sequence upstream of the *fgf14* gene is not found in expanded GAA alleles that cause SCA27B (21). Such protective phenomena could deter the generation of IAs at particular loci, as observed in the current study.

The worldwide allele frequencies of IA might also influence the prevalence of disease in the population, at both the *HTT* and *ATXN1* loci. Though the middle Eastern haplotypes are believed to be of African origin, the prevalence of HD is much lower in Africa. In Alzheimer’s disease, a protective SNP was seen to overcome the risk for disease conferred by ApoeE4 in Africa (22). A focused understanding of the surrounding gene sequences and preserved haplotypes might be needed to uncover such genetic effects in TNRs.

Whole genome data from diverse populations such as India should be examined for occurrence of intermediate alleles in conjunction with other functional DNA repair pathway variants which might influence somatic variation and hence disease presentation. Intermediate alleles that occur alongside pathogenic alleles in the same locus, or at alternate TNR loci (as identified in our study), could also act as disease modifiers in some instances. This kind of data analysis for repeat loci is best carried out using long read sequencing data using increasingly affordable newer technologies, which provide better quality sequence data.

Thus, repeat instability may lead to generation of intermediate alleles in the population which ultimately progress to pathogenic alleles in some tissues in an individual patient. The study of genetic epidemiology of intermediate alleles, its impact on both germline and somatic instability, and correlations with phenotypes, might help explain prevalence of the linked diseases across populations, as well as disease biology.

## Data Availability

All data produced in the present study are available upon reasonable request to the authors

## Notes

**Funding:** This work was supported by grant from Scientific Knowledge for Ageing and Neurological Ailments (SKAN) trust project ‘Genetics of bipolar disorder and related illnesses: a study of intermediate alleles in candidate genes with repeat expansion loci’ (SKAN/002/208/2021/01480), Accelerator program for Discovery in Brain disorders using Stem cells (ADBS) (jointly funded by the Department of Biotechnology, Government of India, and the Pratiksha Trust; Grant BT/PR17316/MED/31/326/2015), and the Centre for Brain and Mind (CBM) grant of the Rohini Nilekani Philanthropies. BV is also supported by The Wellcome Trust DBT India Alliance Intermediate (Clinical and Public Health) Fellowship (IA/CPHI/20/1/505266).

### Competing Interest Statement

The authors have declared no competing interest.

### Funding Statement

This work was supported by grant from Scientific Knowledge for Ageing and Neurological Ailments (SKAN) trust project Genetics of bipolar disorder and related illnesses: a study of intermediate alleles in candidate genes with repeat expansion loci (SKAN/002/208/2021/01480), Accelerator program for Discovery in Brain disorders using Stem cells (ADBS) (jointly funded by the Department of Biotechnology, Government of India, and the Pratiksha Trust; Grant BT/PR17316/MED/31/326/2015), and the Centre for Brain and Mind (CBM) grant of the Rohini Nilekani Philanthropies. BV is also supported by The Wellcome Trust DBT India Alliance Intermediate (Clinical and Public Health) Fellowship (IA/CPHI/20/1/505266).

### Author Declarations

Intitutional ethics committee of National Institute of mental health and neurosciences, India gave ethical approval for this work

