## Supplementary table 1 and 2 for "Intermediate repeat alleles at polyglutamine repeat loci in the Indian population"

**Supplementary Tables:**

Supplementary table 1a: Frequency of intermediate alleles according to Genetic ancestry group

| Population (N) | AFR  (6642) | AMI  (52) | AD AMR  (461) | ASJ  (357) | EAS  (668) | FIN  (1076) | MID EASTERN  (112) | NFE  (6229) | OTHER  (81) | SAS  (648) | GLOBAL  (16326) |
| --- | --- | --- | --- | --- | --- | --- | --- | --- | --- | --- | --- |
| Alleles |  |  |  |  |  |  |  |  |  |  |  |
| ATXN1(36-38) | 238 | 1 | 10 | 14 | 1 | 118 | 1 | 404 | 4 | 22 | 813 |
| ATXN2(32-34) | 11 | 0 | 0 | 0 | 1 | 0 | 0 | 7 | 0 | 0 | 19 |
| ATXN3(45-55) | 4 | 0 | 0 | 0 | 0 | 0 | 0 | 0 | 0 | 0 | 4 |
| HTT(27-35) | 329 | 0 | 39 | 14 | 9 | 36 | 12 | 368 | 10 | 17 | 834 |

Supplementary table 1b: Allele frequency of intermediate alleles at *ATXN1* and *HTT* according to Genetic ancestry group

|  | AFR | AMI | AD AMR | ASJ | EAS | FIN | MID EASTERN | NFE | OTHER | SAS | GLOBAL |
| --- | --- | --- | --- | --- | --- | --- | --- | --- | --- | --- | --- |
| ATXN1 | 0.036 | 0.019 | 0.022 | 0.039 | 0.001 | 0.11 | 0.009 | 0.065 | 0.049 | 0.034 | 0.05 |
| HTT | 0.05 | 0 | 0.085 | 0.039 | 0.013 | 0.033 | 0.107 | 0.059 | 0.123 | 0.026 | 0.051 |

AFR- African/African American, AMI- Amish, AD AMR- Admixed American, ASJ- Ashkenazi Jewish, EAS- East Asians, FIN- European (Finnish), MID- EASTERN- Middle Eastern, NFE – European (Non-Finnish), SAS- South Asian, Global- Average population frequency
